# Nipah virus outbreak in Kerala state, India amidst of COVID-19 pandemic

**DOI:** 10.1101/2021.12.09.21267278

**Authors:** Pragya D. Yadav, Rima R. Sahay, B Anukumar, Sreelekshmy Mohandas, Chandni Radhakrishnan, Mangesh D Gokhale, R Balasubramaniam, Priya Abraham, Nivedita Gupta, AP Sugunan, Rajan Khobragade, Kalpana George, Anita Shete, Savita Patil, Ullas Padinjaremattathil Thankappan, Hitesh Dighe, Jijo Koshy, Vivek Vijay, R Gayathri, P Jayesh Kumar, Asma Rahim, A. Naveen, Sarala Nair, VR Rajendran, V Jayasree, Triparna Majumdar, Rajlaxmi Jain, Prasanth Vishwanathan, Deepak Y. Patil, Abhinendra Kumar, Dimpal A. Nyayanit, Prasad Sarkale, Ashwini Waghmare, Shrikant Baradkar, Pranita Gawande, Poonam Bodake, Kaumudi Kalele, Jyoti Yemul, Sachin Dhaigude, Manjunath Holepannawar, Sanjay Gopale, Ganesh Chopade, Jitendra Narayan, Basavaraj Mathapati, Manoj Kadam, Abhimanyu Kumar, Annasaheb Suryawanshi, Beena Philomina Jose, Saritha Sivadas, NP Akash, TV Vimisha, KV Keerthi

## Abstract

**Background:** We report here a Nipah virus (NiV) outbreak in Kozhikode district of Kerala state, India which had caused fatal encephalitis in an adolescent male and the outbreak response which led to the successful containment of the disease and the related investigations.

**Methods:** Quantitative real-time RT-PCR, ELISA based antibody detection and whole genome sequencing were performed to confirm the Nipah virus infection. Contacts of the index case were traced and isolated based on risk categorization. Bats from the areas near the epicenter of the outbreak were sampled for throat swabs, rectal swabs and blood samples for Nipah virus screening by real time RT-PCR and anti-Nipah virus bat IgG ELISA. Plaque reduction neutralization test was performed for the detection of neutralizing antibodies.

**Results:** Nipah viral RNA and anti-NiV IgG antibodies were detected in the serum of the index case. Rapid establishment of an onsite NiV diagnostic facility and contact tracing helped in quick containment of the outbreak. NiV sequences retrieved from the clinical specimen of the index case formed a sub-cluster with the earlier reported Nipah I genotype sequences from India with more than 95% similarity. Anti-NiV IgG positivity could be detected in 21% of *Pteropus medius* and 37.73% of *Rousettus leschenaultia*. Neutralizing antibodies against NiV could be detected in *P*.*medius*.

**Conclusions:** Stringent surveillance and awareness campaigns needs to be implemented in the area to reduce human-bat interactions and minimize spill over events which can lead to sporadic outbreaks of NiV.

## Introduction

Nipah virus (NiV) causes a highly lethal disease with acute severe encephalitis and acute respiratory distress syndrome in humans. NiV is a *Paramyxovirus* that was identified for the first time during an outbreak of severe encephalitis among the pig-farmers in Malaysia during 1998^1^. The virus is transmitted to humans by direct contact with the respiratory secretions or body fluids of infected animals such as bats and pigs or by consumption of contaminated fruits/palm sap. Both animal-to-human and human-to-human transmission have been documented ^2^.

The disease has been enlisted as a priority disease in the Research and Development blueprint of the World Health Organization (WHO) from the year 2015. South-East Asia is considered as an endemic region for the virus^2^. In Malaysia and Singapore outbreak, humans acquired the infection through the contact with infected pigs^1,3^. Subsequently, an outbreak with encephalitis among human population was observed in Meherpur, Bangladesh during 2001 where the source of infection was traced down to drinking the contaminated raw palm sap or climbing the trees coated with bat excrement^4^. India has witnessed two outbreaks of Nipah virus encephalitis in the eastern state of West Bengal, bordering Bangladesh^5,6^. The first outbreak of NiV was observed among hospital visitors and health workers after coming in contact with a hospitalized patient in Siliguri during January and February 2001^5^. In the year 2007, a second NiV outbreak was reported from Nadia district of West Bengal^6^. A case fatality of 70-100% was observed during these two outbreaks^5,6^. Since 2010, Indian Council of Medical Research-National Institute of Virology (ICMR-NIV), Pune has taken up the surveillance of NiV in bat populations across the country. During this, the presence of NiV was detected among *Pteropus medius* from Myanaguri, West Bengal in 2010 and Cooch Bihar district, West Bengal and Dhubri district, Assam in 2015^7,8^.

After a decade of the last outbreak, a dreadful emergence of NiV was observed in Kozhikode district, Kerala State during May 2018 with case fatality rate of 89%^9^. The outbreak was contained with the quick actions of national and state health system^10^. A year later, a young male from Ernakulam district, Kerala got infected with NiV. Due to the early detection; the further spread of the virus was quickly curtailed. A detailed outbreak investigation to find the source of NiV infection was carried out by ICMR-NIV, Pune during these outbreaks which showed the presence of NiV and anti-NiV antibodies in *Pteropus medius*^11^. The unprecedented emergence of NiV in the Kerala state in 2018 and 2019 raised a serious concern. Recently a NiV outbreak was reported in Kozhikode Kerala in September 2021, where an adolescent male who presented with acute encephalitis and tested positive for NiV, succumbed to the infection.

Here, we describe the 2021 NiV outbreak management in Kozhikode district, Kerala, India with emphasis on the field laboratory set up and quick diagnosis along with the bat survey to trace the source of infection. Immediately after confirming the NiV infection in September 2021, outbreak containment response was initiated in the state of Kerala.

## Materials and Methods

### Case history

In August 2021, an adolescent male (index case) resident of Kozhikode district, Kerala state, India developed low grade fever (Supplementary Figure.1). His family took him to a nearby private clinic (Hospital-1) and sought treatment for fever. In August 2021 his condition deteriorated and he was transferred to another hospital (Hospital-2). In September 2021, the patient’s condition deteriorated further and he developed symptoms of acute encephalitis and myocarditis. On the request of his family, he was transferred to a tertiary care hospital in Kozhikode (Hospital-3). Magnetic resonance imaging of the brain showed multiple small infarcts in the cerebellum, cerebrum, medulla oblongata and pons. With a high suspicion of NiV infection, his clinical samples including plasma, EDTA blood, serum, and cerebrospinal fluid (CSF) samples, endotracheal (ET) secretion and bronchial wash were sent to ICMR-NIV in September 2021.

### Bat trapping and sample collection

For understanding the source of NiV infection and considering a brief history given by parent of the index case regarding the consumption of fruit from the orchard near the house, bat sampling was done during the month of September 2021. The sampling was performed with the prior approval from the Institutional Animal Ethics Committee, Institutional Biosafety Committee of ICMR-NIV, Pune and the Principal Chief Conservator of Forests, Government of Kerala. Four roosting sites were chosen from nearby vicinity of the index case house for sample collection (Supplementary Figure.1). These sites were in Kodiyathur, Cheruvadi, Omassery and Thamarassery) in the state of Kerala, India. Bats were trapped using the mist nets as described earlier^11^. Body weight, sex, secondary sexual characters and forearm length were noted. Blood (n=91), throat (n=102) and rectal swab (n=102) samples were collected from the trapped bats following isoflurane anesthesia. Species identification was performed by mitochondrial Cytochrome b gene PCR as described earlier.^12^

### Real time quantitative RT-PCR

Real-time reverse transcriptase polymerase chain reaction (qRT-PCR) was performed on the samples for the NiV diagnosis as described earlier^13^. One ml of serum/swab samples were used for RNA extraction in an extraction machine using Magmax Viral RNA isolation kit as per manufacturer’s instructions. Five microlitre extracted RNA was used for qRT-PCR using primers designed for Nipah NP gene.^13^ For SARS-CoV-2 detection, qRT-PCR was performed for throat/nasal swab samples using primers for E gene as described earlier^14^.

### Anti-Nipah human IgM ELISA

The assay was performed as described earlier^11^. Briefly, the microtiter plates were coated with 1: 100 diluted anti-human IgM antibodies (Sigma, USA) overnight at 4^0^C. Following blocking of the wells with 2% bovine serum albumin for 2 hours at room temperature, 100 µl of 1:100 diluted heat inactivated serum samples were added to duplicate wells and was incubated at 37°C for one hour. After washing with wash buffer for four times, inactivated NiV antigen (positive antigen) and Vero-CCL81 cell lysate (negative antigen) were added and incubated for one hr at 37°C. The plates were washed and anti-NiV biotinylated antibodies were added and incubated for one hour. This was followed by washing and addition of Streptavidine Horseradish peroxidase (HRP) and incubation for 30 min at 37°C. As substrate, one hundred µl of 3, 3’,5,5’-Tetramethylbenzidine (TMB) was added and incubated for 10 minutes. The reaction was stopped by 1 N sulphuric acid, and the absorbance was measured at 450 nm. The assay cut off were an Optical Density (OD) value more than or equal to 0.20 and the positive/negative (P/N) ratio more than or equal to 1.5.

### Anti-Nipah human IgG ELISA

Briefly, microtiter plates were coated with 0.1 ml of inactivated NiV antigen and normal Vero CCL81 cell lysate as negative antigen in carbonate buffer (pH 9.2, 0.025 M) overnight at 4°C. The wells were blocked with liquid plate sealer (Candor, USA) for 2 hours at room temperature. The plates were washed five times using 10 mM PBS (pH 7.4) with 0.1% Tween-20 (Sigma, USA) at the end of each step. Hundred micro litre of 1:100 diluted heat inactivated human serum samples were added to both positive and negative antigen wells and was incubated at 37°C for one hour. After washing, 100 micro litre of 1:15000 dilution of anti-human IgG HRP antibodies were added and incubated for one hr at 37°C. After washing, 100 µl of TMB substrate was added and incubated for 10 minutes. The reaction was stopped by 1 N sulphuric acid and plates were read at 450 nm. OD value more than or equal to 0.35 and the P/N ratio more than or equal to 1.5 was considered as positive.

### Whole Genome Sequencing (WGS)

To identify the NiV genotype, WGS was carried out on different clinical samples obtained from the patient. RNA was extracted from the clinical samples and WGS was performed using the methods descried earlier^15^. The viral reads generated were analyzed using reference-based mapping, performed in CLC Genomics Workbench version 21.0.4. In order to retrieve the complete genome sequence of the virus all the generated reads were mapped to the reference genome. Phylogenetic analyses and the amino acid variations of the retrieved NiV sequence with the other representative NiV sequences of earlier outbreaks were performed. SARS-CoV-2 positive samples were sequenced as per the method described earlier to identify the lineage in circulation^16^.

### Anti-NiV bat IgG ELISA

The assay was performed as described earlier^15^. NiV antigen/uninfected Vero CCL81 cell lysate antigen coated microtiter plates were used. Diluted bat sera samples were added and incubated for one hour in the microtiter plates at 37°C. The plates were washed four times following the incubation period using wash buffer. Hundred microliter of anti-bat IgG HRP conjugate (1:2000 dilution) was added and incubated. TMB substrate (Clinical Science Product incorporation NeA blue, USA) was added after washing and was again incubated at 37°C for 15 minutes. The reaction was stopped and plates were read at 450 nm.

### Plaque reduction neutralization test

Heat inactivated bat serum samples were mixed with NiV (GenBank accession number: MH523642) containing 50 plaque forming units in 1:1 ratio so as to make a final 10-fold dilution of the serum virus mixture. Anti-Nipah IgG positive mice serum was used as positive control and Anti-Nipah IgG negative mice serum was used as negative control. The mixture was incubated for 1 hour and was added to 24-well tissue culture plate containing a confluent monolayer of Vero CCL-81 cells. The plate was incubated in a CO2 incubator at 37°C for one hour and an overlay medium containing 2% Carboxy methyl cellulose in 2X Minimal Essential Media with 2% fetal bovine serum was added after removing the inoculum. The plate was further incubated at 37°C in CO_2_ incubator for 3 days. After removing the overlay medium, the plate was washed and stained with amido black. The plaques were counted. The titre was defined as the highest serum dilution that resulted in 50 per cent (PRNT50) reduction in the number of plaques.

## Results

### Detection and confirmation of NiV infection

NiV infection was confirmed by qRT-PCR and by the detection of anti-Nipah IgM antibodies in serum sample of the patient. Virus isolation attempts from the samples in Vero CCL-81 cells were not successful. In September 2021, NiV outbreak in Kerala was declared by the Ministry of Health and Family Welfare, Government of India. The patient succumbed to the infection in September 2021. Contact tracing and isolation of high-risk contacts, augmentation of laboratory testing capacity, bat sampling and laboratory investigations were undertaken following the event as described below.

### Establishment of an on-site field NiV diagnostic facility

A team from ICMR-NIV Pune had set up a field diagnostic laboratory in the Department of Microbiology, Government Medical College, Kozhikode, Kerala by September 2021 following all the essential biosafety guidelines and standard operating procedures. The field laboratory screened a total of 125 NiV suspected asymptomatic contacts as well as non-epidemiologically linked suspected NiV cases by qRT-PCR.^10^

### Risk categorization and contact tracing

After declaration of NiV outbreak, systematic field investigations were undertaken to identify the epidemiologically linked close contacts, including health care workers, family members, neighbours, bystanders, etc. The close contacts were classified into primary contacts and secondary contacts and were further grouped into high-risk and low-risk contacts. The high-risk category included individuals with either a history of direct contact with body fluids (blood, urine, saliva, vomitus, etc.) of the confirmed NiV case/a probable case that died without laboratory confirmation or having spent about 12 hour nearby or in closed space with confirmed NiV case. The low-risk contacts were categorized as those having contact with the confirmed NiV case through touching or contact with clothes, linen, or any other items. Samples of symptomatic contacts were shipped to the Biosafety Level 4 (BSL-4) laboratory of ICMR-NIV, Pune for diagnosis. All asymptomatic contacts were tested on site at the field laboratory. Considering the ongoing COVID-19 pandemic, all the close contacts were also screened for SARS-CoV-2.

A total of 240 contacts were listed and among them 64 close contacts [33 female/31 male] were identified and grouped into primary high-risk (n=50) and low-risk (n=9) contacts; secondary high-risk (n=3) and low-risk (n=2) contacts (Figure. 1). Out of 59 primary contacts, 40 were asymptomatic while all the 5 secondary contacts were asymptomatic (Supplementary Table.1). All the 64 close contacts tested negative for NiV by qRTPCR and anti-NiV IgM and IgG ELISA. The throat/nasal swab of the 12 close contacts (symptomatic-8 and asymptomatic-4) were found positive for SARS-CoV-2 by qRT-PCR (Supplementary Table.1). On sequencing of SARS-CoV-2 positive samples (n=12), Delta variant (B.1.167.2) and its derivatives (AY.26) were detected in ten and two cases respectively.

**Figure 1:**
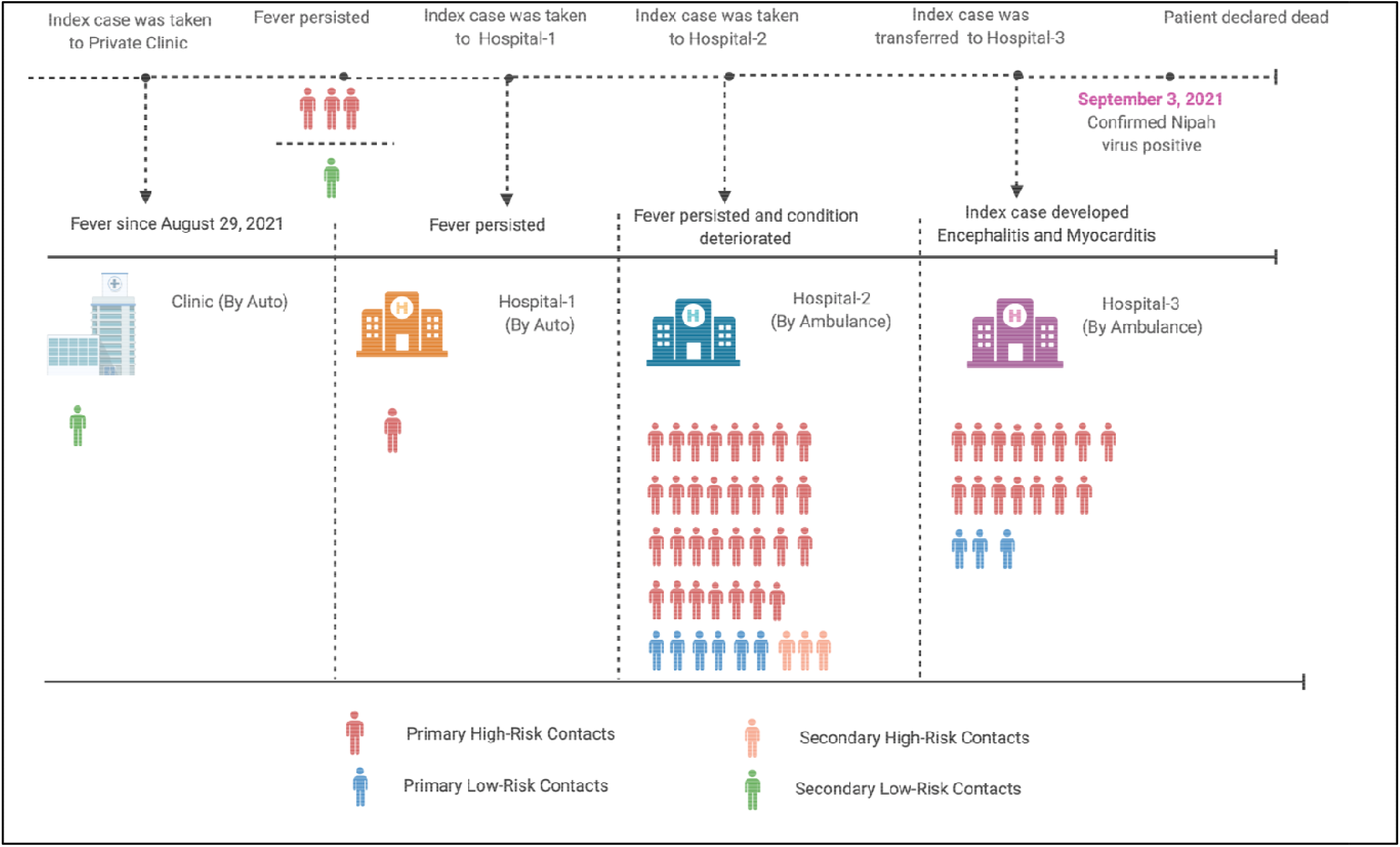
Contact tracing and probable transmission dynamics in the Nipah virus outbreak in Kozhikode district, Kerala state, India, 2021

### Genomic characterization of NiV from clinical specimens of the index case

A phylogenetic analysis was performed for the retrieved NiV sequence (17066 nucleotide) with the other representative NiV sequences of the earlier outbreaks (Figure.2, Supplementary table 2). The retrieved sequence clearly segregated from the Bangladesh NiV sequences and clustered into earlier described Indian (‘I’) genotype. The retrieved sequence showed 99.62 and 99.51 percent nucleotide similarity (PNS) with sequences obtained from human samples during 2018 NiV outbreak and *P. medius* samples during 2019 outbreak respectively. A detailed PNS of the representative NiV sequences with that of the NiV sequence from Malaysia (Accession Number: NC_002728.1) is provided in the Table.1.

**Figure 2:**
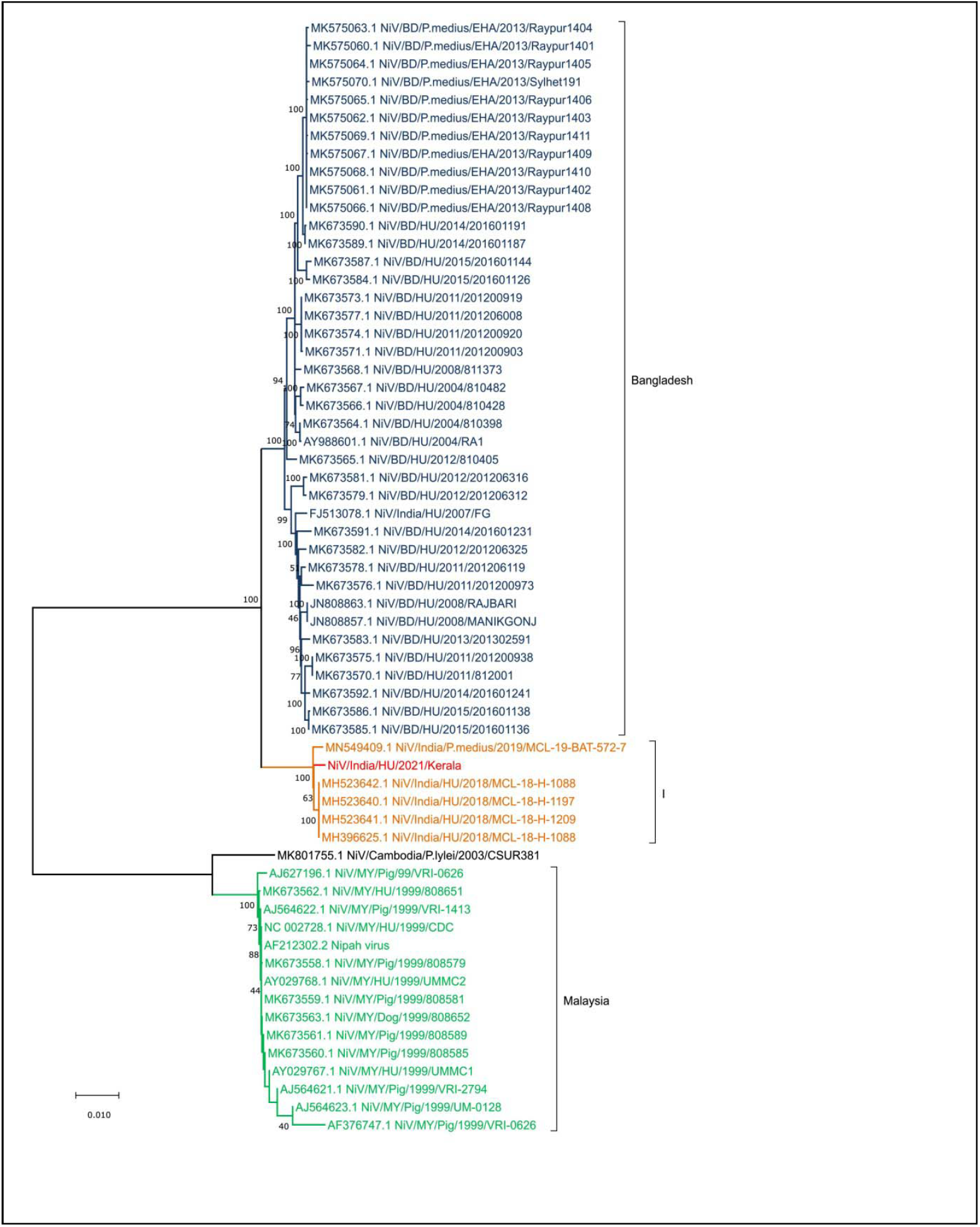
Neighbor Joining tree of Nipah virus full genome obtained from the samples of index case in Kerala outbreak 2021

**Table 1:** PNS for the representative NiV strains retrieved from GenBank database relative to NC_002728.1_NiV/MY/HU/1999/CDC.

| NiV strains | PNS | NiV strains | PNS |
| --- | --- | --- | --- |
| NiV/India/HU/2021/sample | 91.3 | MK673560.1_NiV/MY/Pig/1999/808585 | 100 |
| MN549409.1_NiV/India/P.medius/2019/MCL-19-BAT-572-7 | 91.5 | MK673559.1_NiV/MY/Pig/1999/808581 | 100 |
| MK801755.1_NiV/Cambodia/P.lylei/2003/CSUR381 | 97.7 | MK673558.1_NiV/MY/Pig/1999/808579 | 100 |
| MK673592.1_NiV/BD/HU/2014/201601241 | 91.6 | MK575070.1_NiV/BD/P.medius/EHA/2013/Sylhet191 | 91.6 |
| MK673591.1_NiV/BD/HU/2014/201601231 | 91.6 | MK575069.1_NiV/BD/P.medius/EHA/2013/Raypur1411 | 91.6 |
| MK673590.1_NiV/BD/HU/2014/201601191 | 91.6 | MK575068.1_NiV/BD/P.medius/EHA/2013/Raypur1410 | 91.6 |
| MK673589.1_NiV/BD/HU/2014/201601187 | 91.7 | MK575067.1_NiV/BD/P.medius/EHA/2013/Raypur1409 | 91.6 |
| MK673587.1_NiV/BD/HU/2015/201601144 | 91.5 | MK575066.1_NiV/BD/P.medius/EHA/2013/Raypur1408 | 91.6 |
| MK673586.1_NiV/BD/HU/2015/201601138 | 91.6 | MK575065.1_NiV/BD/P.medius/EHA/2013/Raypur1406 | 91.6 |
| MK673585.1_NiV/BD/HU/2015/201601136 | 91.6 | MK575064.1_NiV/BD/P.medius/EHA/2013/Raypur1405 | 91.6 |
| MK673584.1_NiV/BD/HU/2015/201601126 | 91.6 | MK575063.1_NiV/BD/P.medius/EHA/2013/Raypur1404 | 91.8 |
| MK673583.1_NiV/BD/HU/2013/201302591 | 91.6 | MK575062.1_NiV/BD/P.medius/EHA/2013/Raypur1403 | 91.6 |
| MK673582.1_NiV/BD/HU/2012/201206325 | 91.7 | MK575061.1_NiV/BD/P.medius/EHA/2013/Raypur1402 | 91.6 |
| MK673581.1_NiV/BD/HU/2012/201206316 | 91.7 | MK575060.1_NiV/BD/P.medius/EHA/2013/Raypur1401 | 91.6 |
| MK673579.1_NiV/BD/HU/2012/201206312 | 91.7 | MH523642.1_NiV/India/HU/2018/MCL-18-H-1088 | 91.5 |
| MK673578.1_NiV/BD/HU/2011/201206119 | 91.7 | MH523641.1_NiV/India/HU/2018/MCL-18-H-1209 | 91.5 |
| MK673577.1_NiV/BD/HU/2011/201206008 | 91.7 | MH523640.1_NiV/India/HU/2018/MCL-18-H-1197 | 91.6 |
| MK673576.1_NiV/BD/HU/2011/201200973 | 91.6 | MH396625.1_NiV/India/HU/2018/MCL-18-H-1088 | 91.5 |
| MK673575.1_NiV/BD/HU/2011/201200938 | 91.6 | JN808863.1_NiV/BD/HU/2008/RAJBARI | 91.7 |
| MK673574.1_NiV/BD/HU/2011/201200920 | 91.7 | JN808857.1_NiV/BD/HU/2008/MANIKGONJ | 91.7 |
| MK673573.1_NiV/BD/HU/2011/201200919 | 91.7 | FJ513078.1_NiV/India/HU/2007/FG | 91.6 |
| MK673571.1_NiV/BD/HU/2011/201200903 | 91.7 | AY988601.1_NiV/Bangladesh/HU/2004/RA1 | 91.8 |
| MK673570.1_NiV/BD/HU/2011/812001 | 91.6 | AY029768.1_NiV/MY/HU/1999/UMMC2 | 100 |
| MK673568.1_NiV/BD/HU/2008/811373 | 91.8 | AY029767.1_NiV/MY/HU/1999/UMMC1 | 100 |
| MK673567.1_NiV/BD/HU/2004/810482 | 91.7 | AJ627196.1_NiV/MY/Pig/99/VRI-0626 | 99.7 |
| MK673566.1_NiV/BD/HU/2004/810428 | 91.7 | AJ564623.1_NiV/MY/Pig/1999/UM-0128 | 100 |
| MK673565.1_NiV/BD/HU/2012/810405 | 91.8 | AJ564622.1_NiV/MY/Pig/1999/VRI-1413 | 100 |
| MK673564.1_NiV/BD/HU/2004/810398 | 91.8 | AJ564621.1_NiV/MY/Pig/1999/VRI-2794 | 100 |
| MK673563.1_NiV/MY/Dog/1999/808652 | 100 | AF376747.1_NiV/MY/Pig/1999/VRI-0626 | 99.9 |
| MK673562.1_NiV/MY/HU/1999/808651 | 99.9 | AF212302.2_Nipah_virus | 100 |
| MK673561.1_NiV/MY/Pig/1999/808589 | 100 |  |  |

The different genes of NiV showed amino acid variation between the Bangladesh and I genotype cluster. The changes observed were in the N gene (S503N, P520S, E752G, Q758E, R818H, I820L, T919N, Q982R, A1162T, and G1216D), F gene (I15L) and G gene (R344M, I384V, V427I) of I genotype of the NiV compared to the NC_002728.1. Similarly, the changes were observed in the N gene (R505K, S900G, D921N), G gene (R344K, K386E, T498K) and L gene (R1262K, N 237D) when the Bangladesh (BD) NiV sequences were compared to the NC_002728.1 (Table.2).

**Table.2:**
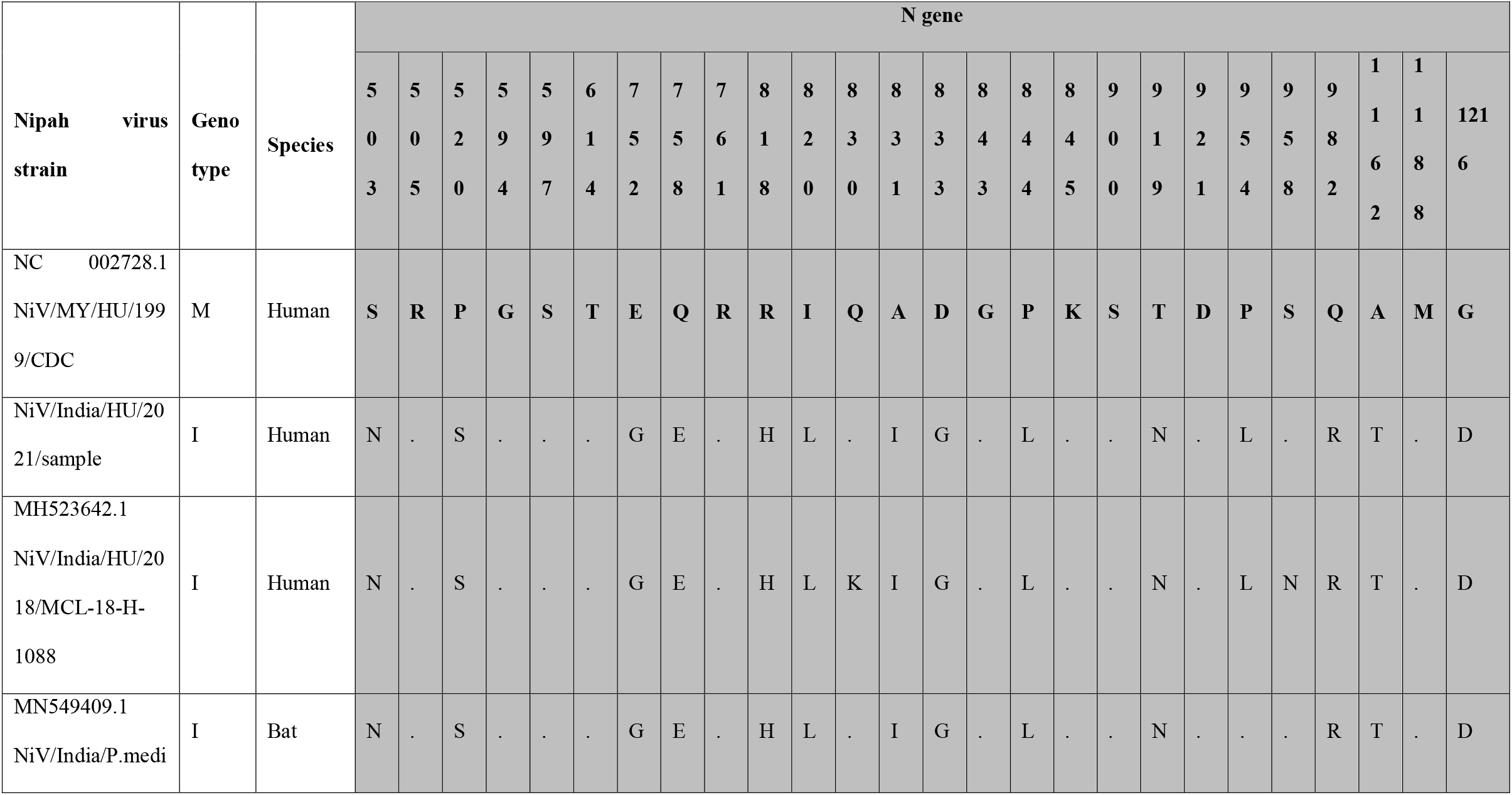
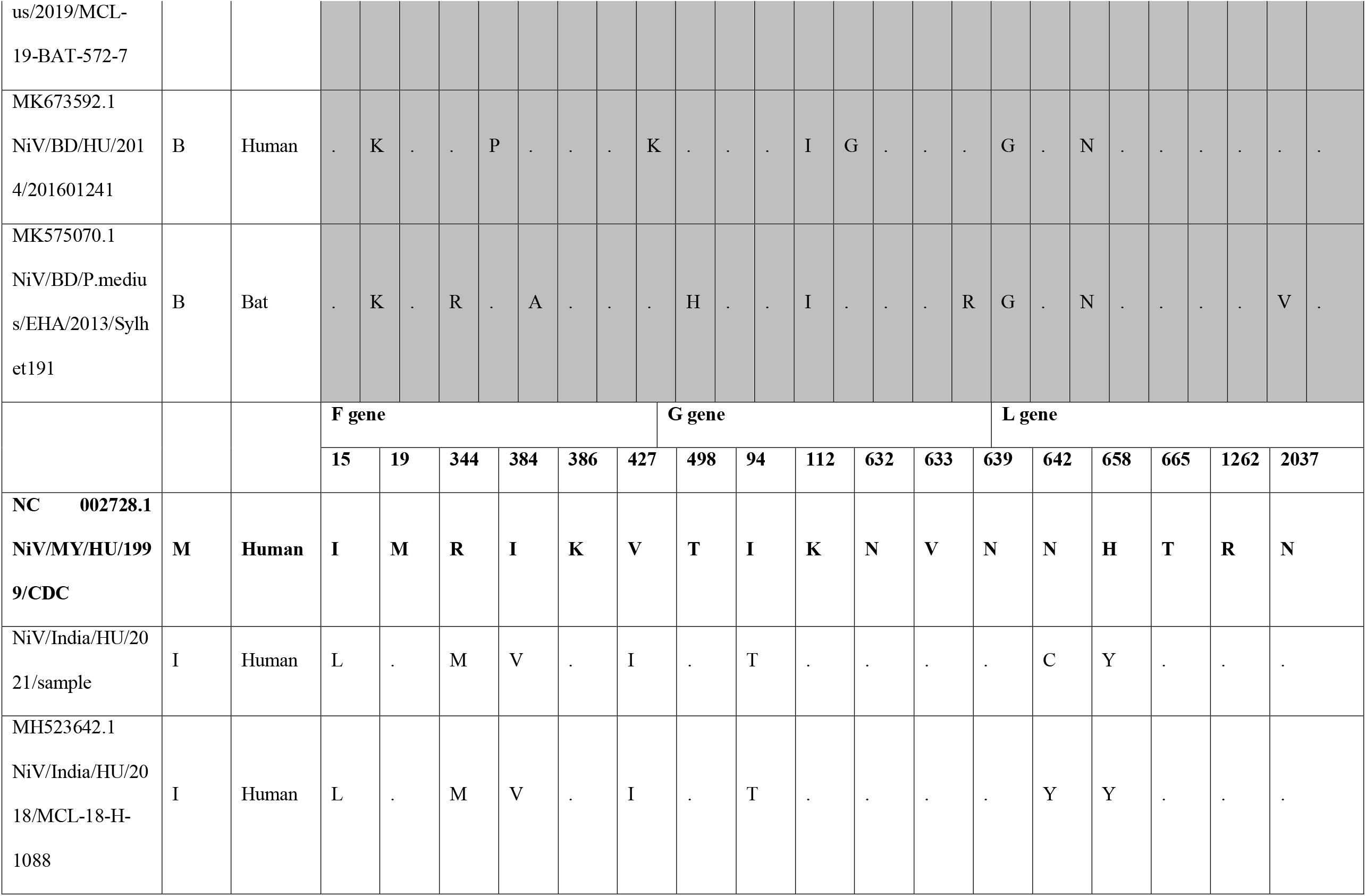
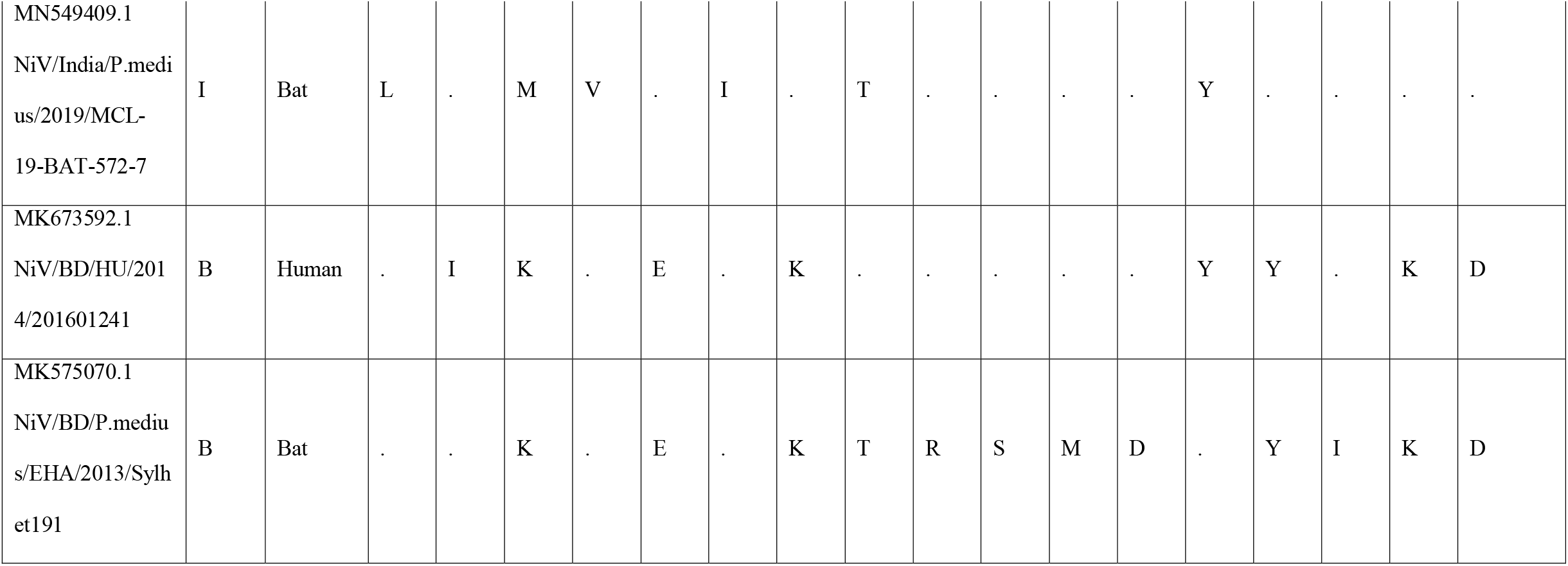
Amino acid changes in the different genotype across representative human and bat sequences for genes encoded by NiV relative to NC 002728.1

### Detection of anti-NiV IgG antibodies in bat samples

The bat species sampled in the study included *Pteropus medius (n=38), Rousettus leschenaultia (n=63)* and *Pipistrellus sp. (n=1)* (Supplementary Table.3). All the bat samples were found to be negative for Nipah viral RNA (Table.3). The serum samples of *P. medius (n=8)* and *R. leschenaultia (n=20)* were tested positive for anti-NiV IgG antibodies. The *P. medius* samples from two sites ie., Kodiyathur and Thamarassery showed positivity of 20% and 56% by ELISA respectively and was further confirmed by PRNT. Two samples were excluded from the assay due to insufficient sample quantity. *R leschenaulti* samples which showed seropositivity by ELISA were found negative for neutralizing antibodies (Table.3).

**Table. 3:** Results and real time RT-PCR, anti-Nipah IgG ELISA and PRNT in bat samples. *2 samples excluded from PRNT assay excluded due to insufficient sample volume

|  | **Bat species** | **Location of capture** | **Real time RT-PCR**<br>**positivity/total tested** | **Anti-Nipah IgG positivity**<br>**by ELISA/total tested**<br>**(%)** | **Anti-Nipah IgG positivity**<br>**by PRNT/total tested** |
| --- | --- | --- | --- | --- | --- |
| 1 | *Pteropus medius* | Kodiyathur | 0/16 | **3/15 (20)** | 3/3 |
|  |  | Cheruvadi | 0/2 | 0/2 (0) | 0/0 |
|  |  | Omassery | 0/12 | 0/12 (0) | 0/0 |
|  |  | Thamarassery | 0/9 | **5/9 (55.6)\*** | 3/3 |
| 2 | *Rousettus leschenaultii* | Kodiyathur | 0/5 | **1/4 (25)** | 0/1 |
|  |  | Cheruvadi | 0/43 | **19/38 (50)** | 0/17 |
|  |  | Omassery | 0/12 | 0/10 | 0/0 |
|  |  | Thamarassery | 0/3 | 0/1 | 0/0 |
| 3 | *Pipistrellus sp.* | Cheruvadi | 0/1 | 0/0 | 0/0 |

## Discussion

Nipah virus outbreaks have been reported from Malaysia, Singapore, Bangladesh and India with a range of clinical presentations and case fatality rate of 40 to 100%.^1-7^ Malaysia and Singapore had reported single NiV outbreak episode whereas Bangladesh reports annual outbreaks of NiV^4^. In India NiV outbreaks have been localized to two regions ie., in the North eastern region of West Bengal and in the southernmost region of Kerala which are separated by a distance of more than 2000 km.^5,6,9-11^ NiV outbreaks are mostly sporadic and the magnitude of the outbreak can be restricted by prompt public health response and containment measures. In 2018, the first NiV outbreak was reported in Kerala state with significant number of losses of lives (91% fatality) and in the year 2019 and 2021 outbreaks in Kerala reported only single case without any further human to human transmission.^9,11^ The diagnosis of the NiV infection is difficult considering the overlapping presentations of the acute respiratory infections and encephalitis syndromes. In the current SARS-CoV-2 pandemic scenario, the diagnosis became even more challenging considering the overlapping clinical features. The quick outbreak containment response and the biosafety practices followed during the COVID-19 pandemic period in Kerala state might have helped in preventing further transmission and restricting the outbreak to a single case.

The role of intermediate hosts like pigs and bats has been demonstrated in the NiV transmission cycle in the previous reported outbreaks.^1,3,4^ *Pteropus* genus of bats appears to be the major reservoir of the NiV.^8,18,20^ *P. medius* is the only *Pteropus* genus bat present in the Indian subcontinent.^19^ Pteropus bats has shown NiV RNA positivity or seropositivity from Indian states like West Bengal, Assam, Haryana and Kerala indicating the risk of spill-over.^7,8,11,20^ Viral genome recovered from the current outbreak also clustered with the previously reported NiV genotype from Kerala in bats and humans samples. This suggests a stable genotype circulating locally in bat population in Kerala. Research has shown NiV evolves at a slower rate compared to other similar RNA viruses.^21,22^ In 2018, the outbreak occurred in Perambra, Kozhikode which is about 40 km far from the location of the present outbreak.^9^ A total of 23 cases and 21 deaths were reported. 21% of the *Pteropus medius* bats surveyed in the vicinity of the index case residence. In 2019, NiV RNA positivity was documented in *P medius* bats from Thodupuzha which is more than 200 km from Kozhikode district.^11^ *Pteropus* species movement could also play an important role in virus spread, but in India such records are not available. Home ranges of *P. medius* appear to be smaller than *P. vampyrus*, another frugivorous bat in Malaysia and this depend on food availability^18^. Studies on the *Pteropus* species movement and connectivity among the bat populations could help us in understanding the potential of virus spread to bat colonies of adjacent areas.

NiV RNA could not be detected in any of the bats samples in the present study. Previous studies have also reported a low PCR positivity in *P medius*. ^18,23,24^. Virus shedding in bats is driven by multiple factors like individual immune status, pregnancy, virus recrudescence, stress etc.^18,24^ Re-infection in adult bats are possible in approximately 7 years^24^. We detected 25% and 3.2% NiV positivity in bats from outbreak regions of Kerala in 2018 and 2019.^9,11^ High seroprevalence in the bat colonies can dampen the virus transmission.^18^ Seasonality of NiV outbreaks have been reported is linked to the breeding season and fruit harvesting season.^25^ Unlike the previous reported outbreaks in the month of May and June in Kerala, the present outbreak was in August end. A cross sectional spatial study conducted between 2006 and 2012 in Bangladesh reports the ability of NiV shedding by *P. medius* throughout the year.^18^

During the current outbreak investigations, we could detect antibodies in a total of 21% *P. medius* bats. If we further classify this positivity, site wise, a 20% positivity was observed in Kodiyathur, a site within 1 km distance from the index case house and 55.6% in Thamarassery, a site 18 km far from this outbreak. During the investigation, 2 juvenile *P. medius* bats were found positive for anti-NiV IgG antibodies. The seroprevalence in the juveniles is an indicator of enzootic cycle of NiV in recent past. Waning of immunity with time and loss of maternal antibodies could affect the transmission dynamics within the bat colonies.^18,24^ Local NiV epizootics in bats contribute to the outbreaks in Bangladesh^.18^ Sporadic natures of outbreaks in case of NiV are explained by the NiV dynamics in the resident bat colonies and the bat-human interface resulting in spill over.

Non-pteropid bats have shown NiV seropositivity in many countries.^17,23,26,27^ In India, other than *P medius*, bats like *R. leschenaulti*a and *P. pipistrellu*s have shown seropositivity by ELISA.^17^ Interspecies transmission could be possible with other frugivorous bats as they share same habitat for food resources. Similarly, in the present study, we could detect seropositivity in *R. leschenaulti*a bats by ELISA. But absence of NAb against NiV in these bats points to the possibility of detection of related Henipaviruses in these bats. Similar observations are reported in Vietnam where the ELISA positive samples of *R. leschenaulti*a failed to show any NAb, indicating the probability of cross reacting antibodies against non-neutralizing epitopes^26^. Antibody positivity by ELISA and Western Blot in *R leschenaulti*a has been reported from China, but neutralization studies were not performed in the samples^27^.

The findings from the current outbreak, suggest that spill-over of NiV infection in humans is sporadic and the seroprevalence in bats indicates the prevalence of the NiV infections in the *P. medius* population. Stringent surveillance measures and awareness campaigns to limit interactions with bats needs to be implemented in the area as the NiV transmission dynamics depends on multiple host factors including human behavior and human-bat interface. For early detection and containment of NiV outbreaks, it is critical to strengthen human surveillance for Acute Encephalitis Syndrome and Severe Acute Respiratory Infection, including testing for NiV in susceptible areas.

## Data Availability

All data produced in the present study are available upon reasonable request to the authors

## Conflict of Interest

The authors declare that the research was conducted in the absence of any commercial or financial relationships that could be construed as a potential conflict of interest.

## Funding

ICMR supported the funding for this study under the mission-oriented project to the ICMR-National Institute of Virology, Pune. The funders had no role in study design, data collection or interpretation, or the decision to submit the work for publication. The findings and conclusions in this study are of the authors.

## Acknowledgments

Authors extend gratitude to Veena George [Hon’ble Minister for Health and Social Justice, Kerala], for her exemplary leadership and efficient coordination of the Nipah virus disease control activities, and “The Team Kerala Health,” the district administration, press, media, and people for their efforts in streamlining the public health responses. The authors are thankful to Prof. Balram Bhargava, Director General, ICMR and Dr Samiran Panda, Scientist G & Head, Epidemiology and Communicable Diseases Division, ICMR for their support. Authors are also thankful for the staff of the Administration section, ICMR-NIV Pune for their support during the outbreak response. We also acknowledge the help and support received from Dr. Arun Sacharia, Chief Veterinarian (Wild life) and Dr. Arun Sathyan, Assistant Veterinarian, Thamaraserry during bat survey.

## Notes

### Competing Interest Statement

The authors have declared no competing interest.

### Author Declarations

Approved by Institutional Ethics Committee, Institutional Biosafety Committee of Indian Council of Medical Research - National Institute of Virology, Pune and the Principal Chief Conservator of Forests, Government of Kerala.

