## Supplementary File for "Nipah virus outbreak in Kerala state, India amidst of COVID-19 pandemic"

1.1 Supplementary Figures

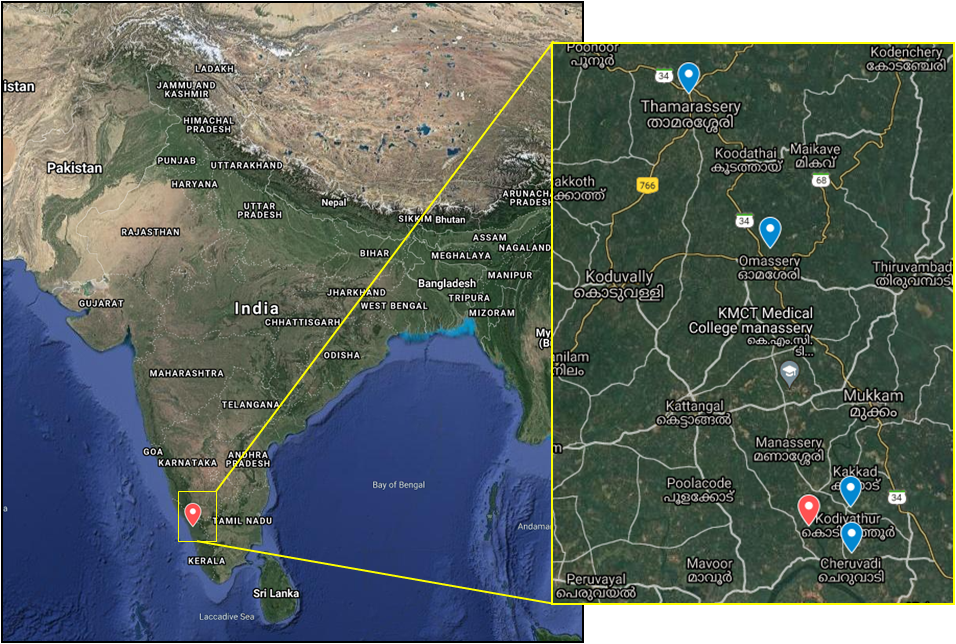

Supplementary Figure 1: The map showing the Nipah virus outbreak location in India and in inset, the location of index case house (red icon) and the four bat sampling sites (blue icon).

### 1.2 Supplementary Tables

**Supplementary Table 1: Details of the Close contacts screened during Nipah outbreak in Kozhikode district, Kerala state, 2021**

| **Sr. No** | **Laboratory ID** | **Sex** | **Primary/ Secondary Contacts** | **High-Risk /Low-Risk Contacts** | **Location of contact** | **Relationship with Index Case** | **Symptomatic/ Asymptomatic** | **If symptomatic, then symptoms details** | **Nipah Real time RTPCR** | **Anti-Nipah IgM and Anti-Nipah IgG Antibodies** | **SARS-CoV-2 Real time RT-PCR** |
| --- | --- | --- | --- | --- | --- | --- | --- | --- | --- | --- | --- |
| 1 | MCL-21-H-9728 | Male | Primary | High-Risk | Household | Close relative | Symptomatic | Cold, Rhinitis and Myalgia | Negative | Negative | Positive |
| 2 | MCL-21-H-9731-1 | Female | Primary | High-Risk | Household | Close relative | Symptomatic | Cold, Rhinitis and Myalgia | Negative | Negative | Positive |
| 3 | MCL-21-H-9755-1 | Male | Primary | High-Risk | Household | Close relative | Symptomatic | Cold, Rhinitis and Myalgia | Negative | Negative | Positive |
| 4 | MCL-21-H-9746 | Male | Primary | High-Risk | Hospital-3 | HCW | Symptomatic | Cold, Rhinitis and Myalgia | Negative | Negative | Positive |
| 5 | MCL-21-H-9743 | Female | Primary | High-Risk | Hospital-3 | HCW | Symptomatic | Cold, Rhinitis and Myalgia | Negative | Negative | Negative |
| 6 | KZ-NIV-37-TS | Male | Primary | High-Risk | Hospital-2 | HCW | Asymptomatic | NA | Negative | Negative | Negative |
| 7 | MCL-21-H-9749 | Female | Primary | High-Risk | Hospital-2 | HCW | Symptomatic | Cold, Rhinitis and Myalgia | Negative | Negative | Negative |
| 8 | KZ-NIV-31-TS | Male | Primary | High-Risk | Hospital-2 | HCW | Asymptomatic | NA | Negative | Negative | Negative |
| 9 | KZ-NIV-21-TS | Female | Primary | High-Risk | Hospital-3 | HCW | Asymptomatic | NA | Negative | Negative | Negative |
| 10 | KZ-NIV-25-TS | Male | Primary | High-Risk | Hospital-2 | HCW | Asymptomatic | NA | Negative | Negative | Negative |
| 11 | KZ-NIV-23-TS | Male | Primary | High-Risk | Hospital-2 | HCW | Asymptomatic | NA | Negative | Negative | Negative |
| 12 | KZ-NIV-36-TS | Female | Primary | High-Risk | Hospital-3 | HCW | Asymptomatic | NA | Negative | Negative | Negative |
| 13 | KZ-NIV-26-TS | Female | Primary | High-Risk | Hospital-3 | HCW | Asymptomatic | NA | Negative | Negative | Negative |
| 14 | KZ-NIV-24-TS | Female | Primary | High-Risk | Hospital-3 | HCW | Asymptomatic | NA | Negative | Negative | Negative |
| 15 | KZ-NIV-27-TS | Female | Primary | High-Risk | Hospital-3 | HCW | Asymptomatic | NA | Negative | Negative | Negative |
| 16 | KZ-NIV-14-TS | Female | Primary | High-Risk | Hospital-3 | HCW | Asymptomatic | NA | Negative | Negative | Negative |
| 17 | KZ-NIV-17-TS | Female | Primary | High-Risk | Hospital-3 | HCW | Asymptomatic | NA | Negative | Negative | Negative |
| 18 | KZ-NIV-32-TS | Female | Primary | High-Risk | Hospital-3 | HCW | Asymptomatic | NA | Negative | Negative | Negative |
| 19 | KZ-NIV-29-TS | Male | Primary | High-Risk | Hospital-3 | HCW | Asymptomatic | NA | Negative | Negative | Negative |
| 20 | MCL-21-H-9737 | Female | Primary | High-Risk | Hospital-2 | Patient | Symptomatic | Cold, Rhinitis and Myalgia | Negative | Negative | Negative |
| 21 | KZ-NIV-06-TS | Male | Primary | High-Risk | Hospital-2 | Patient | Asymptomatic | NA | Negative | Negative | Negative |
| 22 | KZ-NIV-09-TS | Male | Primary | High-Risk | Hospital-2 | Patient | Symptomatic | Cold, Rhinitis and Myalgia | Negative | Negative | Positive |
| 23 | MCL-21-H-9740 | Female | Primary | High-Risk | Hospital-2 | Patient | Symptomatic | Cold, Rhinitis and Myalgia | Negative | Negative | Negative |
| 24 | KZ-NIV-12-TS | Female | Primary | High-Risk | Hospital-2 | Bystander | Asymptomatic | NA | Negative | Negative | Negative |
| 25 | KZ-NIV-07-TS | Male | Primary | High-Risk | Hospital-2 | Bystander | Asymptomatic | NA | Negative | Negative | Negative |
| 26 | KZ-NIV-05-TS | Female | Primary | High-Risk | Hospital-2 | Bystander | Asymptomatic | NA | Negative | Negative | Negative |
| 27 | KZ-NIV-08-TS | Male | Primary | High-Risk | Hospital-2 | Bystander | Asymptomatic | NA | Negative | Negative | Negative |
| 28 | KZ-NIV-03-TS | Male | Primary | High-Risk | Hospital-2 | Bystander | Asymptomatic | NA | Negative | Negative | Negative |
| 29 | MCL-21-H-9761 | Female | Primary | High-Risk | Hospital-3 | HCW | Symptomatic | Cold, Rhinitis and Myalgia | Negative | Negative | Positive |
| 30 | KZ-NIV-33-TS | Female | Primary | High-Risk | Hospital-2 | HCW | Asymptomatic | NA | Negative | Negative | Negative |
| 31 | KZ-NIV-34-TS | Female | Primary | High-Risk | Hospital-2 | HCW | Asymptomatic | NA | Negative | Negative | Negative |
| 32 | KZ-NIV-28-TS | Female | Primary | High-Risk | Hospital-2 | HCW | Asymptomatic | NA | Negative | Negative | Negative |
| 33 | KZ-NIV-18-TS | Male | Primary | High-Risk | Hospital-2 | HCW | Asymptomatic | NA | Negative | Negative | Negative |
| 34 | KZ-NIV-30-TS | Female | Primary | High-Risk | Hospital-2 | HCW | Asymptomatic | NA | Negative | Negative | Negative |
| 35 | KZ-NIV-15-TS | Female | Primary | High-Risk | Hospital-2 | HCW | Asymptomatic | NA | Negative | Negative | Negative |
| 36 | KZ-NIV-35-TS | Male | Primary | High-Risk | Hospital-2 | HCW | Asymptomatic | NA | Negative | Negative | Negative |
| 37 | KZ-NIV-22-TS | Female | Primary | High-Risk | Hospital-2 | HCW | Asymptomatic | NA | Negative | Negative | Positive |
| 38 | KZ-NIV-20-TS | Female | Primary | High-Risk | Hospital-3 | HCW | Asymptomatic | NA | Negative | Negative | Negative |
| 39 | KZ-NIV-40-TS | Male | Primary | High-Risk | Hospital-2 | HCW | Asymptomatic | NA | Negative | Negative | Positive |
| 40 | KZ-NIV-41-TS | Male | Primary | High-Risk | Hospital-2 | HCW | Asymptomatic | NA | Negative | Negative | Negative |
| 41 | KZ-NIV-45-TS | Male | Primary | High-Risk | Hospital-2 | Patient | Asymptomatic | NA | Negative | Negative | Negative |
| 42 | MCL-21-H-9798 | Female | Primary | High-Risk | Hospital-3 | HCW | Symptomatic | Cold, Rhinitis and Myalgia | Negative | Negative | Negative |
| 43 | KZ-NIV-48-TS | Female | Primary | High-Risk | Hospital-3 | HCW | Asymptomatic | NA | Negative | Negative | Negative |
| 44 | KZ-NIV-46-TS | Male | Primary | High-Risk | Hospital-2 | Volunteer | Asymptomatic | NA | Negative | Negative | Negative |
| 45 | MCL-21-H-9789 | Female | Primary | High-Risk | Hospital-2 | HCW | Symptomatic | Cold, Rhinitis and Myalgia | Negative | Negative | Negative |
| 46 | MCL-21-H-9801 | Female | Primary | High-Risk | Hospital-1 | HCW | Symptomatic | Cold, Rhinitis and Myalgia | Negative | Negative | Positive |
| 47 | MCL-21-H-9804 | Male | Primary | High-Risk | Hospital-2 | Patient | Symptomatic | Cold, Rhinitis and Myalgia | Negative | Negative | Negative |
| 48 | MCL-21-H-9813 | Female | Primary | High-Risk | Hospital-2 | HCW | Symptomatic | Cold, Rhinitis and Myalgia | Negative | Negative | Negative |
| 49 | MCL-21-H-9816 | Female | Primary | High-Risk | Hospital-2 | HCW | Symptomatic | Cold, Rhinitis and Myalgia | Negative | Negative | Negative |
| 50 | MCL-21-H-9984 | Female | Primary | High-Risk | Hospital-2 | HCW | Symptomatic | Cold, Rhinitis and Myalgia | Negative | Negative | Negative |
| 51 | KZ-NIV-16-TS | Male | Primary | Low-Risk | Hospital-3 | HCW | Asymptomatic | NA | Negative | Negative | Negative |
| 52 | KZ-NIV-19-TS | Male | Primary | Low-Risk | Hospital-2 | HCW | Asymptomatic | NA | Negative | Negative | Positive |
| 53 | KZ-NIV-44-TS | Female | Primary | Low-Risk | Hospital-3 | HCW | Asymptomatic | NA | Negative | Negative | Negative |
| 54 | KZ-NIV-39-TS | Male | Primary | Low-Risk | Hospital-2 | Patient | Asymptomatic | NA | Negative | Negative | Negative |
| 55 | KZ-NIV-42-TS | Male | Primary | Low-Risk | Hospital-2 | HCW | Asymptomatic | NA | Negative | Negative | Negative |
| 56 | KZ-NIV-47-TS | Female | Primary | Low-Risk | Hospital-2 | HCW | Asymptomatic | NA | Negative | Negative | Negative |
| 57 | MCL-21-H-9795 | Female | Primary | Low-Risk | Hospital-2 | HCW | Symptomatic | Cold, Rhinitis and Myalgia | Negative | Negative | Negative |
| 58 | KZ-NIV-38-TS | Male | Primary | Low-Risk | Hospital-2 | HCW | Asymptomatic | NA | Negative | Negative | Negative |
| 59 | MCL-21-H-9831 | Male | Primary | Low-Risk | Hospital-3 | HCW | Symptomatic | Cold, Rhinitis and Myalgia | Negative | Negative | Negative |
| 60 | KZ-NIV-11-TS | Male | Secondary | High-Risk | Hospital-2 | Bystander | Asymptomatic | NA | Negative | Negative | Negative |
| 61 | KZ-NIV-04-TS | Male | Secondary | High-Risk | Hospital-2 | Bystander | Asymptomatic | NA | Negative | Negative | Positive |
| 62 | KZ-NIV-10-TS | Male | Secondary | High-Risk | Hospital-2 | Bystander | Asymptomatic | NA | Negative | Negative | Negative |
| 63 | MCL-21-H-9758 | Male | Secondary | Low-Risk | Private Clinic | Auto driver- commuted the patient to Clinic from Home | Symptomatic | Cold, Rhinitis and Myalgia | Negative | Negative | Positive |
| 64 | MCL-21-H-9792 | Male | Secondary | Low-Risk | Household | Family Member | Symptomatic | Cold, Rhinitis and Myalgia | Negative | Negative | Negative |

**Supplementary table 2**: Percent read mapped and genome recovered from the clinical samples of index case during Kerala outbreak 2021.

| **Sl no.** | **Specimen** | **Total reads** | **Relevant reads** | **Genome length** | **% genome recovered** |
| --- | --- | --- | --- | --- | --- |
| 1 | Cerebrospinal Fluid | 3,892,234 | 2,792,957 | 14434 | 79.08174 |
| 2 | Plasma | 3,095,834 | 99,835 | 13561 | 74.29871 |
| 3 | Bronchial wash | 8,814,670 | 30,387 | 15767 | 86.38505 |
| 4 | Endotracheal secretion | 3,164,422 | 16,502 | 13164 | 72.1236 |
| 5 | Whole blood | 7,274,278 | 39,013 | 6062 | 33.2128 |
| 6 | Citrate Plasma | 4,854,126 | 26,446 | 4629 | 25.3616 |

**Supplementary table 3: The details of bats samples during the NiV outbreak, Kozhikode Kerala 2021**

| **Sl no.** | **Bat species** | **Number** | **Sex** | **Body weight in grams**  **(Mean ± SD)** | **Average forearm length in centimeters**  **(Mean ± SD)** |
| --- | --- | --- | --- | --- | --- |
| **1** | *Pteropus medius* | 39 | Male  (n= 23) | 568.34±211.8 | 16.0±1.60 |
|  |  |  | Female  (n=15) | 563.17±179.88 | 15.99±1.09 |
| **2** | *Rousettus leschenaultii* | 62 | Male  (n=20) | 64.07±21.72 | 7.47±0.93 |
|  |  |  | Female  (n=43) | 54.89±14.04 | 7.48±0.63 |
| **3** | *Pipistrellus sp.* | 1 | Male  (n=1) | 3.91 | 2.5 |
